# SARS-CoV-2 infections elicit higher levels of original antigenic sin antibodies compared to SARS-CoV-2 mRNA vaccinations

**DOI:** 10.1101/2021.09.30.21264363

**Authors:** Elizabeth M. Anderson, Theresa Eilola, Eileen Goodwin, Marcus J. Bolton, Sigrid Gouma, Rishi R. Goel, Mark M. Painter, Sokratis A. Apostolidis, Divij Mathew, Debora Dunbar, Danielle Fiore, Amanda Brock, JoEllen Weaver, John S. Millar, Stephanie DerOhannessian, The UPenn COVID Processing Unit, Allison R. Greenplate, Ian Frank, Daniel J. Rader, E. John Wherry, Scott E. Hensley

## Abstract

Severe acute respiratory syndrome coronavirus 2 (SARS-CoV-2) mRNA vaccines elicit higher levels of antibodies compared to natural SARS-CoV-2 infections in most individuals; however, the specificities of antibodies elicited by vaccination versus infection remain incompletely understood. Here, we characterized the magnitude and specificity of SARS-CoV-2 spike-reactive antibodies from 10 acutely infected health care workers and 23 participants who received mRNA-based SARS-CoV-2 vaccines. We found that infection and primary mRNA vaccination elicited S1 and S2-reactive antibodies, while secondary vaccination boosted mostly S1 antibodies. Using magnetic bead-based absorption assays, we found that SARS-CoV-2 infections elicited a large proportion of original antigenic sin-like antibodies that bound efficiently to common seasonal human coronaviruses but poorly to SARS-CoV-2. In converse, vaccination only modestly boosted antibodies reactive to common seasonal human coronaviruses and these antibodies bound efficiently to SARS-CoV-2. Our data indicate that SARS-CoV-2 mRNA vaccinations elicit fundamentally different antibody responses compared to SARS-CoV-2 infections.

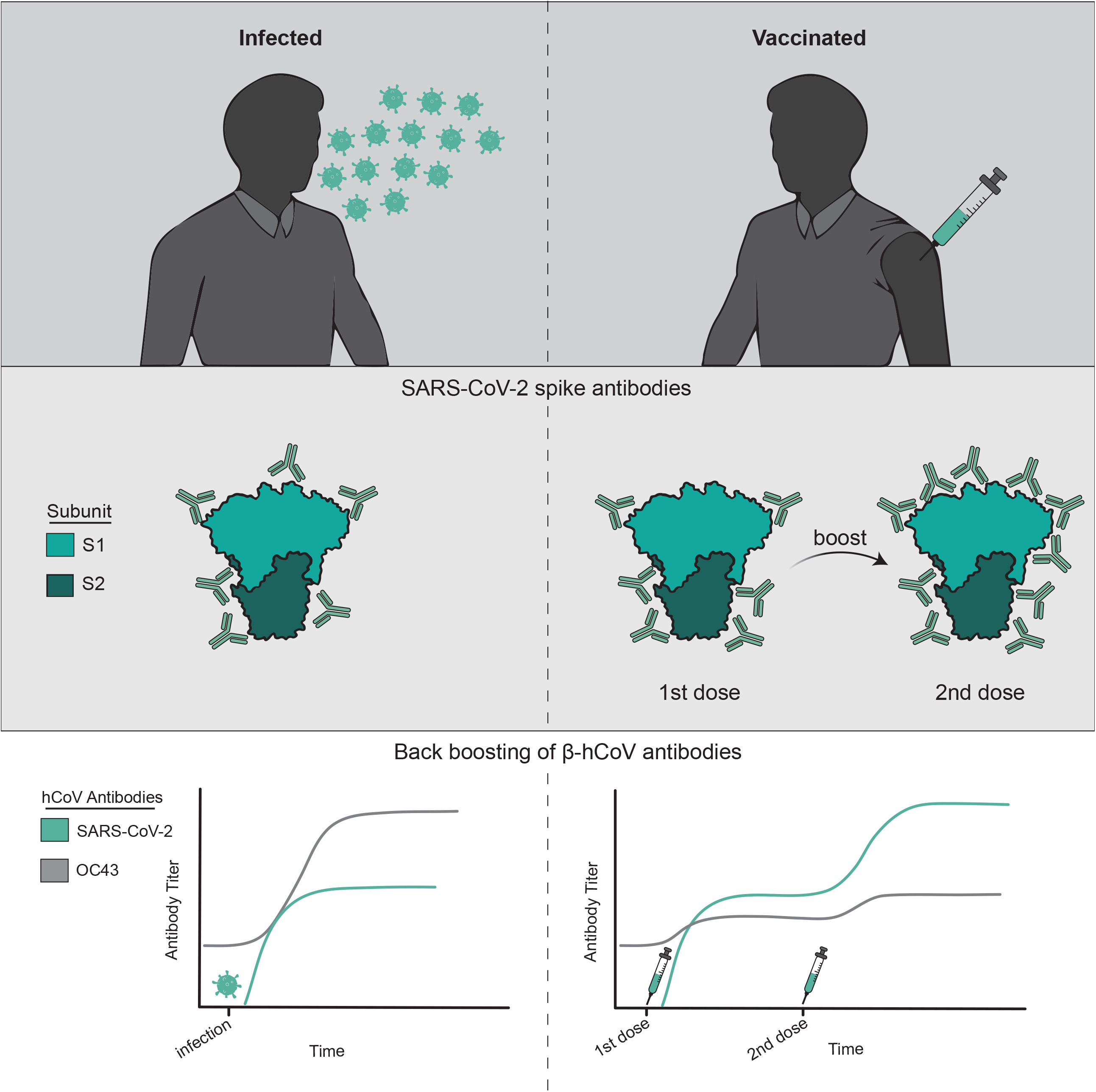

**HIGHLIGHTS:**

- SARS-CoV-2 mRNA vaccines elicit higher levels of antibodies compared to SARS-CoV-2 infections
- The first dose of an mRNA vaccine generates both S1 and S2 responses while the second dose boosts primarily S1-specific antibodies
- SARS-CoV-2 infections, but not mRNA vaccinations, elicit high levels of antibodies that bind strongly to seasonal coronaviruses but weakly to SARS-CoV-2

## BACKGROUND

Since late 2019 severe acute respiratory syndrome coronavirus 2 (SARS-CoV-2) has spread across the world causing a global pandemic^1,2^. This prompted the rapid development of several SARS-CoV-2 vaccines including two that use an mRNA-based platform (reviewed in^3-5^). The mRNA vaccines, Pfizer BNT162b2 and Moderna mRNA-1273, employ lipid nanoparticles that encase modified mRNA encoding the spike protein of SARS-CoV-2^6^. SARS-CoV-2 mRNA vaccines have been found to be safe and effective at preventing severe COVID-19, hospitalizations, and death^7,8^. Recent studies demonstrate that SARS-CoV-2 mRNA vaccines elicit long-lived antibody responses that partially recognize and protect against antigenically distinct SARS-CoV-2 variants^9-15^.

Some studies suggest that prior infections with common seasonal human coronaviruses (hCoVs) impact the severity of SARS-CoV-2 infections^16,17^. Most individuals are exposed to hCoVs early in childhood^18-23^ and then re-exposed to antigenically drifted forms of these viruses throughout life^24-26^. Common hCoVs include the HKU1 and OC43 betacoronaviruses (β-hCoVs) and 229E and NL63 alphacoronaviruses^27-30^. Studies from our group and others have shown that some individuals possessed antibodies that could bind to SARS-CoV-2 proteins prior to the COVID-19 pandemic^18,31,32^. SARS-CoV-2 is a β-hCoV, and antibodies reactive to the OC43 β-hCoV can be boosted upon SARS-CoV-2 infections^16,18,32,33^ and SARS-CoV-2 mRNA vaccinations^34-36^. It is unknown if the recall of β-hCoV antibodies upon SARS-CoV-2 infections impacts disease outcome. A recent study suggests that the recall of OC43 β-hCoV antibodies is associated with a compromised *de novo* SARS-CoV-2 response in individuals with fatal COVID-19^37^.

The boosting of hCoV antibodies upon infection with the antigenically distinct SARS-CoV-2 is consistent with the doctrine of ‘original antigenic sin’, first proposed to describe influenza virus antibody responses by Thomas Francis in 1960^38^. We recently developed new absorption assays to show that sequential heterosubtypic influenza virus infections elicit antibodies that paradoxically do not bind effectively to the boosting viral strain^39^. In the current report, we used a similar absorption technique to define the specificity of β-hCoV antibodies elicited by SARS-CoV-2 infections. We completed a series of studies to determine if these boosted β-hCoV antibodies could cross-react to SARS-CoV-2 and we compared antibodies elicited by SARS-CoV-2 infections and SARS-CoV-2 mRNA vaccinations.

## RESULTS

### SARS-CoV-2 infections and vaccinations elicit antibodies against the SARS-CoV-2 spike protein

We obtained samples from individuals before and after acute SARS-CoV-2 infections (n=10) and pre-/post-two doses of a Pfizer BNT162b2 mRNA SARS-CoV-2 vaccine (n=23). Samples from SARS-CoV-2 infected individuals were obtained from a health care worker sero-monitoring study in which all infections were relatively mild^16^. All 10 individuals were seronegative in the beginning of this study and acquired a PCR-confirmed SARS-CoV-2 infection over the course of the study. Blood samples were collected from SARS-CoV-2 infected individuals 5-28 days (mean 17.9 days) after PCR-confirmed infections. Blood samples were collected from SARS-CoV-2 mRNA vaccinated individuals^40^ 7-15 days post-primary immunization (mean 14.2 days), the day of or day before the booster immunization (mean 21.2 days post-primary), and 7-12 days post-booster immunization (mean 7.5 days post-boost). Individuals in our infection and vaccination studies had similar ages and both groups were predominantly female (**Supplemental Table 1**).

Consistent with previous studies (reviewed in^41^), antibodies against the full-length spike (FL-S) of SARS-CoV-2 increased following infection (**Figure 1A**) and vaccination (**Figure 1B**). SARS-CoV-2 FL-S antibody levels were similar following infections and primary vaccinations, and antibody levels were significantly boosted following the second dose of vaccine (**Figure 1B-C**). The SARS-CoV-2 FL-S protein consists of two domains, the S1 domain which encompasses the receptor binding domain (RBD) essential for cell attachment and entry^42-45^ and the N terminal domain (NTD), and the S2 domain which shares more sequence homology with hCoVs^28,33^. Infection and primary vaccination elicited high levels of both S1 and S2 antibodies (**Figure 1A-B**), whereas secondary vaccinations boosted S1 antibodies more efficiently compared to S2 antibodies (**Figure 1B, D-E**).

**Figure 1.**
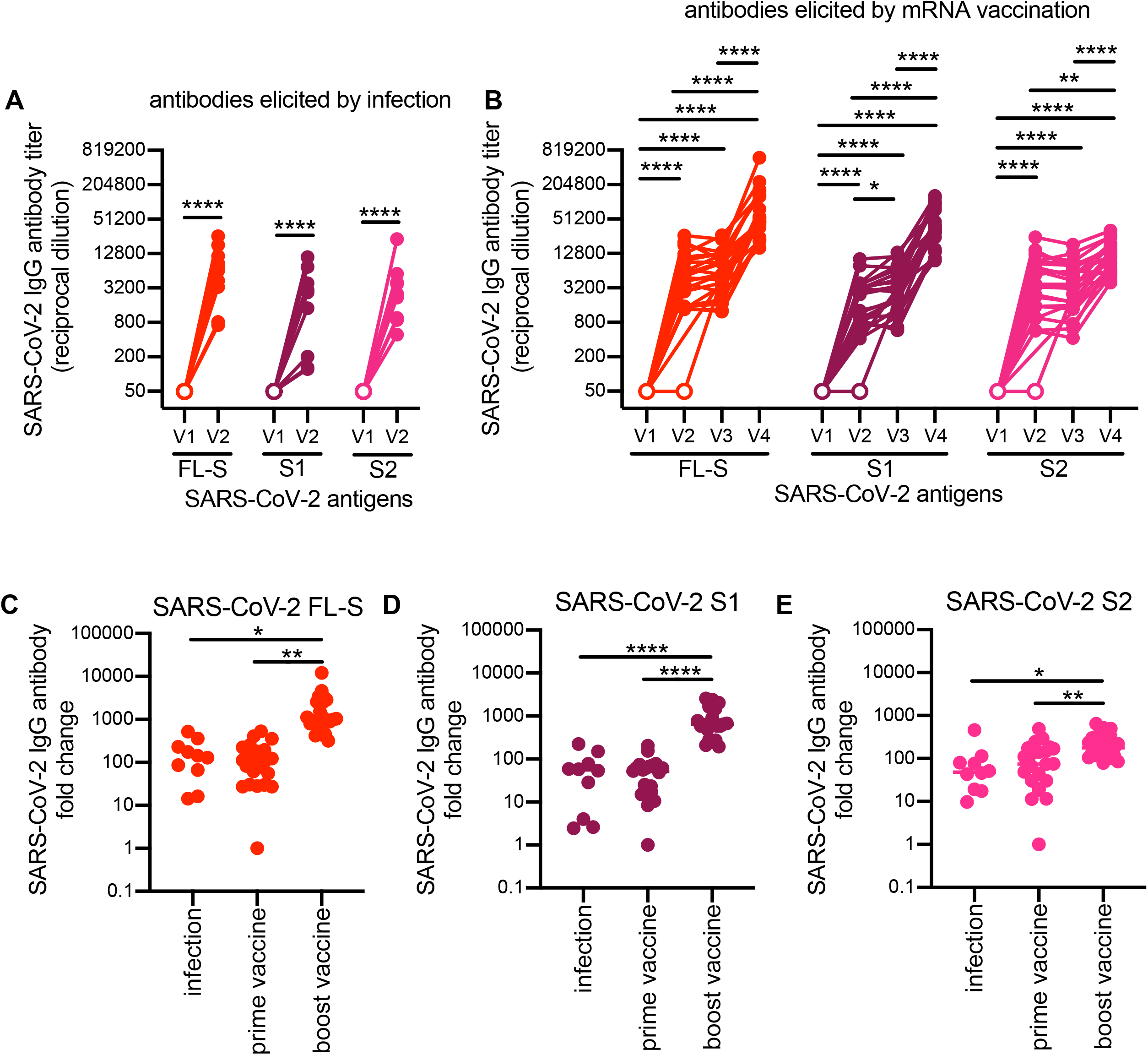
Specificity of SARS-CoV-2 antibodies induced after SARS-CoV-2 infection versus vaccination. ELISAs were completed to quantify levels of serum antibodies binding to the SARS-CoV-2 full-length spike (FL-S) protein, the S1 domain (S1) of S, and the S2 domain (S2) of S after SARS-CoV-2 infection (**A)** and mRNA vaccination (**B**). Paired t-tests of log2 transformed antibody titers ****p<0.0001, ***p<0.001, **p<0.01, *p<0.05. We calculated fold-change in antibody titers before and after seroconversion and pre-/post-prime and boost doses of a SARS-CoV-2 mRNA vaccine (**C-E**). One way ANOVA of antibody fold change ****p<0.0001, **p<0.01 *p<0.05. Horizontal lines indicate geometric mean and error bars represent standard deviation.

### SARS-CoV-2 infections and vaccinations elicit antibodies against a common human betacoronavirus

We previously found that antibodies against the FL-S of the OC43 β-hCoV are boosted upon SARS-CoV-2 infection in hospitalized patients with severe disease^18^. Consistent with these findings, we found that antibodies reactive to the OC43 FL-S increased upon SARS-CoV-2 infections in health care workers with mild disease (**Figure 2A**). A recent study showed that antibody titers against β-hCoVs increase upon SARS-CoV-2 mRNA vaccination^36^. Although we found small increases in OC43 FL-S antibody titers following SARS-CoV-2 mRNA vaccinations (**Figure 2B**), the magnitude of OC43 FL-S antibody boosts were much lower following vaccinations compared to infections for most individuals (**Figure 2C**). Antibodies boosted by infections and vaccinations primarily targeted the S2 domain of the OC43 spike (**Figure 2A-B, 2D-E**), with infections boosting S2 responses more effectively compared to vaccinations (**Figure 2E**). Booster vaccinations did not affect levels of OC43 FL-S, S1, or S2 antibody levels (**Figure 2B-E**).

**Figure 2.**
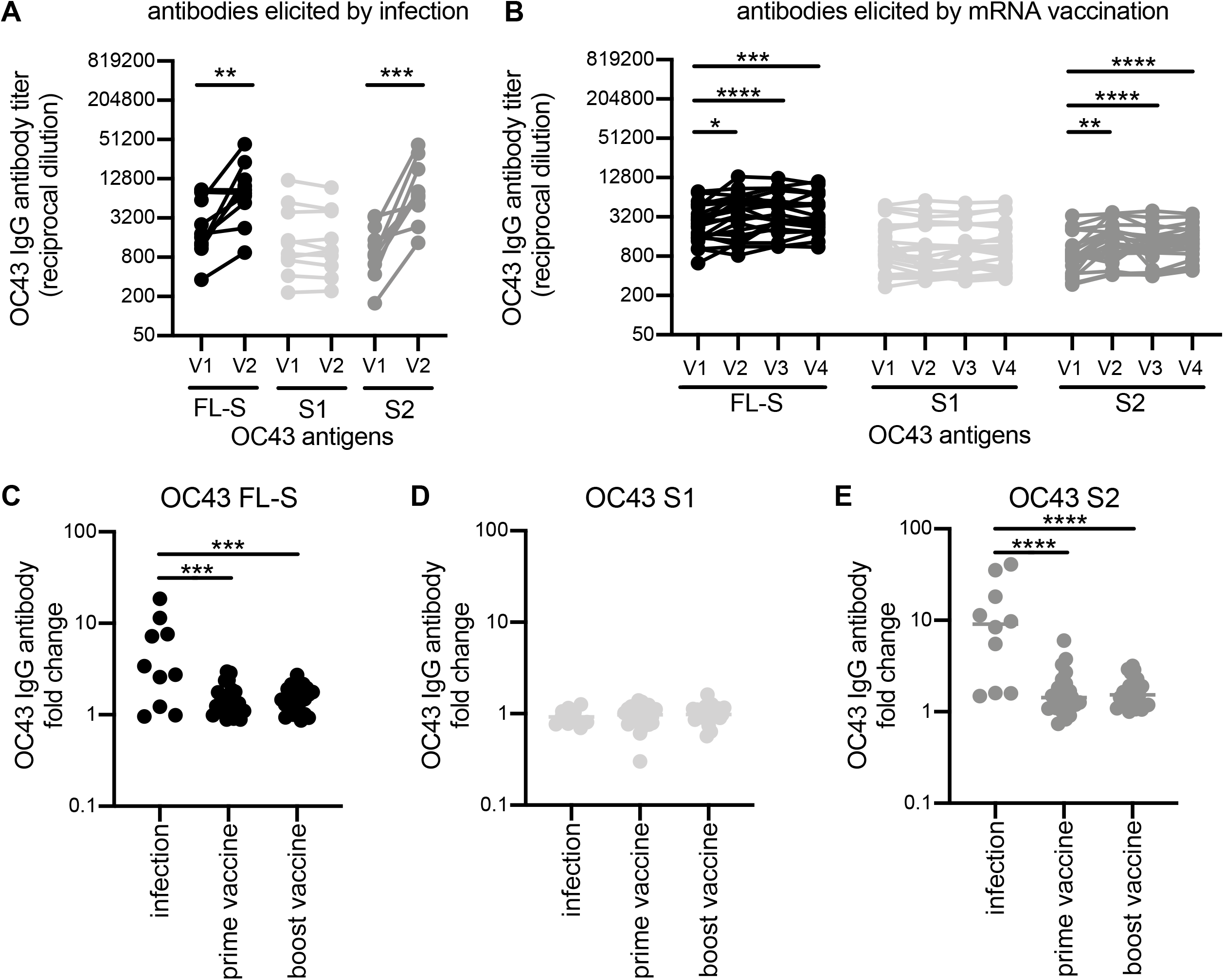
Antibodies to a related seasonal coronavirus are boosted upon SARS-CoV-2 infection and after vaccination to a lesser extent. ELISAs were completed to quantify levels of serum antibodies binding to the betacoronavirus, OC43 full-length spike (FL-S) protein, the S1 domain (S1) of S, and the S2 domain (S2) of S after SARS-CoV-2 infection (**A)** and mRNA vaccination **(B)**. Paired t-tests of log2 transformed antibody titers ****p<0.0001, ***p<0.001, **p<0.01, *p<0.05. We calculated fold-change in antibody titers before and after seroconversion and pre-/post- prime and boost doses of a SARS-CoV-2 mRNA vaccine (**C-E**). One way ANOVA of antibody fold change ****p<0.0001 and ***p=0.0001. Horizontal lines indicate geometric mean and error bars represent standard deviation.

### SARS-CoV-2 infections elicit higher levels of original antigenic sin antibodies compared to SARS-CoV-2 mRNA vaccinations

We previously developed absorption-based assays to measure the level of cross-reactivity of antibodies elicited by influenza virus infections^39^. In that study, we sequentially infected ferrets with two antigenically distinct influenza virus strains and analyzed serum samples collected after each infection. We found that many antibodies elicited by secondary influenza virus infections paradoxically did not bind effectively to the secondary boosting influenza virus strain. We proposed that antigenically distinct influenza viruses engage memory B cells elicited by prior infections through multiple low affinity interactions with thousands of B cell receptors on memory B cells. Low affinity antibodies secreted in a soluble form through this recall response fail to bind to the antigenically distinct recall antigens because they require the level of multivalent binding that is provided on B cells.

We developed a magnetic bead-based absorption assay to determine if OC43 FL-S antibodies elicited by SARS-CoV-2 infection and vaccination cross-react with SARS-CoV-2 FL-S. We incubated serum samples with beads coupled with either OC43 FL-S or SARS-CoV-2 FL-S and then we depleted bead-reactive antibodies using a magnetic column. Then, we quantified antibody levels in serum absorbed with antigen coupled beads to assess antibody cross-reactivity. SARS-CoV-2 FL-S-reactive antibodies elicited by infection were efficiently depleted with SARS-CoV-2 FL-S coupled beads but not OC43 FL-S coupled beads (**Figure 3A**). OC43 FL-S-reactive antibodies boosted by SARS-CoV-2 infection were efficiently depleted with OC43 FL-S coupled beads, but surprisingly these antibodies were not depleted with SARS-CoV-2 FL-S coupled beads (**Figure 3B**). These results are surprising since these OC43 FL-S-reactive antibodies were boosted upon SARS-CoV-2 infection. Vaccination-elicited antibodies had different cross-reactive properties compared to infection-elicited antibodies. Similar to antibodies elicited by infection, SARS-CoV-2 FL-S-reactive antibodies elicited by vaccination were efficiently depleted with SARS-CoV-2 FL-S coupled beads but not OC43 FL-S coupled beads (**Figure 3D**). In contrast to antibodies elicited by infection, OC43 FL-S-reactive antibodies boosted by SARS-CoV-2 vaccination were efficiently depleted with both OC43 FL-S and SARS-CoV-2 FL-S coupled beads (**Figure 3B**). As a control, we quantified levels of influenza virus hemagglutinin (HA) antibodies in these experiments and we found that HA-reactive antibodies were not depleted with SARS-CoV-2 FL-S or OC43 FL-S labeled beads (**Figure 3C, 3F**). Taken together, our data suggest that SARS-CoV-2 infection elicits OC43 FL-S-reactive antibodies that bind poorly to SARS-CoV-2 FL-S, while SARS-CoV-2 mRNA vaccinations elicits a relatively lower level of OC43 FL-S-reactive antibodies that efficiently cross-react with both OC43 FL-S and SARS-CoV-2 FL-S.

**Figure 3.**
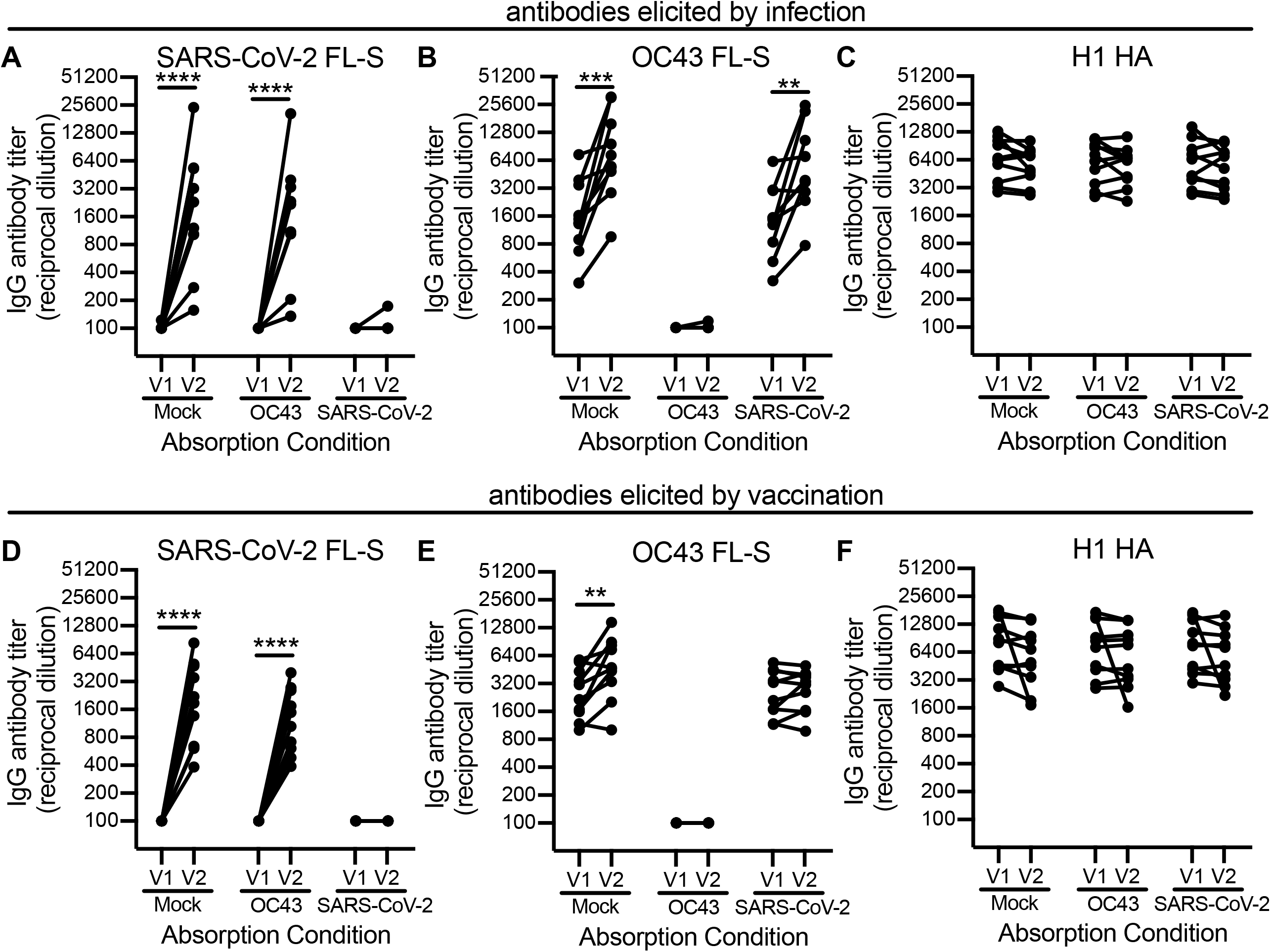
Seasonal coronavirus spike antibodies boosted by SARS-CoV-2 infection do not bind well to SARS-CoV-2 spike. Sera samples from 10 SARS-CoV-2 infected health care workers (**A-C**) and 10 SARS-CoV-2 mRNA vaccinated participants (**D-F**) were absorbed with SARS-CoV-2 full length (FL) S-coupled beads, OC43 Full length (FL) S-coupled beads, or mock treated beads prior to antibody quantification by ELISA. We determined reciprocal antibody titers in samples before and after infection and pre-/post- the first dose of an mRNA vaccine for SARS-CoV-2 FL-S (**A**,**D**), OC43 FL-S (**B**,**E**), and an unrelated antigen, influenza hemagglutinin H1 (**C, F**). Paired t-tests of log2 transformed antibody titers ****p<0.0001, ***p<0.001, **p<0.01.

## DISCUSSION

Here we found that polyclonal antibodies elicited by SARS-CoV-2 mRNA vaccines are at a higher magnitude and have different specificities compared to those elicited by SARS-CoV-2 infections. We demonstrate that primary vaccinations elicit antibodies that bind to the S1 and S2 region of SARS-CoV-2 spike, and that S1-specific antibody responses are preferentially boosted after second vaccine doses. These data are consistent with recent reports that indicate that mRNA vaccinations elicit broader and more diverse antibody responses that are more effective at neutralizing SARS-CoV-2 variants compared to antibodies elicited by infection^15,46-48 49^.

We found major differences in OC43 spike binding between antibodies elicited by SARS-CoV-2 infections versus mRNA vaccinations. SARS-CoV-2 infections elicited high levels of antibodies that bound to the S2 region of the OC43 spike protein; however, our absorption assays demonstrated that these antibodies bound poorly to the SARS-CoV-2 spike. Conversely, we found that SARS-CoV-2 mRNA vaccinations elicited lower levels of antibodies that reacted to the S2 region of the OC43 spike protein. Unlike antibodies elicited by infections, these vaccine-elicited antibodies were truly cross-reactive and bound efficiently to both SARS-CoV-2 and OC43 spike proteins. Further studies will be required to fully understand mechanisms that lead to different types of antibody responses elicited by SARS-CoV-2 infections versus vaccinations. It is possible that memory B cells elicited by prior β-hCoV infections are recalled by both SARS-CoV-2 infections and vaccinations, and that long-lived germinal centers elicited by mRNA vaccinations^50^ are required to allow for somatic hypermutations that promote the formation of cross-reactive S2 antibodies that bind efficiently to the spike proteins of both β-hCoVs and SARS-CoV-2. Consistent with this, a recent study found that S2-specific B cells with a memory phenotype are quickly recruited following primary immunization of humans^51^

Boosting of OC43 S2-reactive antibodies following SARS-CoV-2 infection is consistent with Thomas Francis’ doctrine of ‘original antigenic sin’^38^. Francis found that antibodies elicited by influenza vaccines often bound strongly to influenza virus strains that an individual was exposed to in childhood, although it is not apparent if these recalled influenza virus antibody responses typically occur at the expense of producing *de novo* antibodies (as we reviewed here^52^) The functional consequences of recalling low affinity S2-reactive antibodies following SARS-CoV-2 infections are unclear. Our previous studies found no correlation between OC43-reactive antibody induction and disease outcome following SARS-CoV-2 infection^18^ but a recent study suggested that the recall of OC43-reactive antibodies is associated with a compromised *de novo* SARS-CoV-2 response in individuals with fatal COVID-19^37^. Further studies are required to determine how the induction of different types of hCoV and SARS-CoV-2 antibodies affect disease outcome following SARS-CoV-2 infections.

## STAR METHODS

### KEY RESOURCES TABLE

| REAGENT or RESOURCE | SOURCE | IDENTIFIER |
| --- | --- | --- |
| **Antibodies** |  |  |
| Goat anti-human IgG-HRP | Jackson ImmunoResearch | 109-036-098 |
| mAb CR3022 | Produced for this paper | N/A |
| mAb CR9114 | Produced for this paper | N/A |
| **Biological Samples** |  |  |
| Acutely SARS-CoV-2 infected HCW serum samples | The University of Pennsylvania | Gouma et al., 2021 |
| SARS-CoV-2 mRNA vaccinated participants serum samples | The University of Pennsylvania | Goel et al., 2021 |
| **Chemicals, Peptides, and Recombinant Proteins** |  |  |
| SARS-CoV-2 spike protein | Produced for this paper | N/A |
| SARS-CoV-2 S1 subunit protein | ACROBiosystems | Cat. S1N-C52H3 |
| SARS-CoV-2 S2 subunit protein | ACROBiosystems | Cat. S2N-C52H5 |
| OC43 spike protein | Sino Biological | Cat. 40607-V08B |
| OC43 S1 subunit protein | Produced for this paper | N/A |
| OC43 S2 subunit protein | Produced for this paper | N/A |
| IVR-190 H1N1 rHA | Produced for this paper | N/A |
| **Experimental Models: Cell Lines** |  |  |
| 293T | ATCC | Cat. CRL-3216, RRID:CVCL_0063 |
| 293F | Laboratory of Scott Hensley, University of Pennsylvania, PA | Thermo Fisher cat. R79007 |
| **Recombinant DNA** |  |  |
| Plasmid: pCAGGS SARS-CoV-2 spike | Laboratory of Florian Krammer, Mt. Sinai, NY | Amanat et al., 2020 |
| Plasmid: OC43 rS1 | Laboratory of Scott Hensley, University of Pennsylvania, PA | Anderson et al., 2021 |
| Plasmid: OC43 rS2 | Laboratory of Scott Hensley, University of Pennsylvania, PA | Anderson et al. 2021 |
| Plasmid: mAb CR3022 HC | Laboratory of Ian Wilson, Scripps Research Institute, CA | Yaun et al., 2020 |
| Plasmid: mAb CR3022 LC | Laboratory of Ian Wilson, Scripps Research Institute, CA | Yaun et al., 2020 |
| Plasmid: mAb CR9114 HC | Laboratory of Ian Wilson, Scripps Research Institute, CA | Dreyfus et al., 2012 |
| Plasmid: mAb CR9114 LC | Laboratory of Ian Wilson, Scripps Research Institute, CA | Dreyfus et al., 2012 |
| **Software and Algorithms** |  |  |
| Prism8 | GraphPad Software | www.graphpad.com/scientific-software/prism/ |

## RESOURCES AVAILABILITY

### Lead Contact

Further information and requests for resources and reagents should be directed to and will be fulfilled by the Lead Contact, Scott E. Hensley.

### Materials Availability

All unique reagents generated in this study will be available from the Lead Contact upon reasonable request.

### Data and Code Availability

All raw data generated in this study will be deposited on Mendeley Data:

## EXPERIMENTAL MODEL AND SUBJECT DETAILS

### Samples from Human Subjects

The infection cohort described in this report consists of health care workers within the University of Pennsylvania Healthcare System who were recruited into a SARS-CoV-2 sero-monitoring study that included biweekly blood draws as previously described^16^. A nasal pharyngeal (NP) swab was collected from all health care workers who tested positive for SARS-CoV-2 IgG and or IgM antibodies or who were experiencing COVID-like symptoms during the study period. NP swabs were PCR tested for the presence of SARS-CoV-2 viral RNA. Seroconverted health care workers with PCR-confirmed SARS-CoV-2 infection (n=10 adults ≥18 years old) were included in this analysis.

The vaccination cohort described in this report consists of participants (n=23 adults ≥18 years old) who enrolled in a study at the University of Pennsylvania that included blood draws before and after two vaccination doses with an mRNA-based COVID-19 vaccine as previously described^40^. Whole blood was collected from participants who provided proof of vaccination with Pfizer (BNT162b2) mRNA vaccines. Samples were collected at 4 timepoints: 1-2 weeks before vaccination (baseline) (visit 1; V1), 1-2 weeks post-primary immunization (visit 2; V2), the day of or day before booster immunization (visit 3; V3), and 1-2 weeks post-booster immunization (visit 4; V4). Only participants without prior SARS-CoV-2 exposure were included in this report. Plasma and PBMCs were isolated from whole blood for downstream assays.

All sera and plasma samples were heat-inactivated in a 56^°^C water bath for 1 hour prior to serological testing. All samples were collected after obtaining informed consent and studies were approved by the University of Pennsylvania Institutional Review Board.

### Cell lines

293F cells were from Thermo fisher (Thermo Fisher cat. R79007). 293T cells were from ATCC (ATCC cat. CRL-3216, RRID:CVCL_0063). All cell lines were cultured using manufacturer’s guidelines and used as described in Method Details below.

## METHOD DETAILS

### Proteins for serological studies

SARS-CoV-2 full length spike (FL-S) protein was purified by Ni-NTA resin from 293F cells transfected with a plasmid that encodes the FL-S (A gift from Florian Krammer, Icahn School of Medicine at Mt. Sinai, New York City NY)^53^. S1 and S2 subunits of the SARS-CoV-2 spike were purchased from Acro Biosystems (ACROBiosystems, Newark, DE; cat. S1N-C52H3, and S2N-C52H5, respectively) and reconstituted in 200 μL Dulbecco’s phosphate buffered saline (DPBS) to a final concentration of 500 μg/mL. OC43 FL-S was also purchased (Sino Biological, Wayne PA; cat. 40588-V08B) and reconstituted in DPBS. OC43 subunit proteins were purified in our laboratory as previously described^18^. Briefly, mammalian expression plasmids encoding the S1 (amino acids 15-760) or S2 (amino acids 766-1305) domains of the OC43 spike protein were cloned with an N-terminal OC43 S signal peptide, and a C-terminus encoding a Factor Xa cleavage site, a trimerization domain from T4 fibritin (Foldon), a site-specific biotinylation sequence (AviTag), and a hexa-histidine purification tag. 293F cells were transfected with S1 or S2 encoding plasmids and proteins were purified from cell culture supernatant 6 days later with Ni-NTA resin (Qiagen, Hilden, Germany). Proteins were concentrated and buffer exchanged into PBS with Amicon centrifugal filters (Millipore, Burlington, MA) prior to quantification on a spectrophotometer (NanoDrop, Thermo Fisher Scientific, Waltham, MA).

### ELISAs

Antibodies reactive to SARS-CoV-2 and OC43 antigens were quantified by enzyme-linked immunosorbent assays (ELISA) as previously described^18,54^. Absorbed sera samples were also tested for the presence of influenza virus H1 HA antibodies. In brief, ELISA plates (Thermo Fisher Scientific, Waltham, MA: cat. 14-245-153) were coated overnight at 4^°^C with either 2 μg/mL SARS-CoV-2 or influenza HA antigens, 1.5 μg/mL OC43 antigens, or Dulbecco’s phosphate buffered saline (DPBS) to control for background antibody binding. Sera was heat-inactivated in a 56^°^C water bath for 1 hour prior to serial dilutions starting at 1:50 in dilution buffer. ELISA plates were blocked for 1 hour before 50 μL of diluted sera was added and plates were incubated for 2 hours on an orbital shaker. ELISA plates were washed 3 times with 1x PBS supplemented with 2% Tween (PBS-T) before the addition of goat anti-human IgG conjugated to horseradish peroxidase secondary antibody at a 1:5000 dilution (Jackson ImmunoResearch Laboratories, West Grove, PA: cat. 109-036-098). ELISA plates were developed with TMB substrate, and the reactions were stopped after 5 minutes by the addition of 250 mM hydrochloric acid prior to reading on a SpectraMax 190 microplate reader (Molecular Devices, San Jose, CA). Serum antibody titers were obtained from a standard curve of either serially diluted monoclonal antibody (CR3022 for SARS-CoV-2 or CR9114 for influenza virus HA starting at 0.5μg/mL) or serially diluted pooled serum (for OC43 ELISAs). Standard curves were included on every plate to control for plate-to-plate variation. Antibody titers for each sample were measured in at least two technical replicates performed on separate days.

### Carboxyl magnetic bead absorptions

SARS-CoV-2 FL-S or OC43 FL-S antigens were coupled to carboxyl magnetic beads (RayBiotech, Peachtree Corners, GA; cat. 801-114-2) at a concentration of 35 μg antigen/100 μL magnetic beads. Mock beads were prepared by the addition of DPBS in place of antigen. Briefly, 175 μg of diluted antigen or PBS (for mock) was added to 500 μL of magnetic beads and the mixture was incubated for 2 hours at 4^°^C with constant mixing. The unbound fractions were removed using a magnetic stand and conjugated beads were quenched by the addition of 300 μL 50 mM Tris, pH 7.4 prior to a 15-minute incubation at room temperature with constant mixing. Quenching buffer was removed using a magnetic stand and the conjugated beads were washed 4 times with 300 μL wash buffer (DPBS supplemented with 0.1% BSA and 0.05% Tween-20). After the final wash, beads were resuspended in 300 μL wash buffer and were stored at 4^°^C prior to use in serum absorption assays.

Sera samples were absorbed with beads coupled to SARS-CoV-2 FL-S, OC43 FL-S, and mock beads. Sera samples were diluted in PBS to a final dilution of 1:25. Next, 20 μL of antigen coupled-magnetic beads or mock-treated beads were added to 100 μL of diluted sera and the mixtures were incubated for 1 hour at room temperature on a plate mixer at 800rpm. Fractions containing the unabsorbed antibodies were removed using a 96-well plate magnetic stand. Unabsorbed fractions were diluted in buffer (DPBS supplemented with 1% milk and 0.1% Tween-20) prior to running in ELISA.

## Data Availability

All data are included in the manuscript.

## QUANTIFICATION AND STATISTICAL ANALYSIS

Statistical analyses were performed using Prism version 8 (GraphPad Software, San Diego CA). Reciprocal serum dilution antibody titers were log2 transformed for statistical analysis. ELISA antibody titers below the limit of detection (LOD) were set to a reciprocal titer equal to half the LOD. Log2 transformed antibody titers were compared with paired and unpaired t-tests, and one-way ANOVAs with Tukey’s multiple comparisons. Statistical significance was defined as a p-value <0.05.

## ACKNOWLEDGEMENTS

This project has been funded in part with Federal funds from the National Institute of Allergy and Infectious Diseases, National Institutes of Health, Department of Health and Human Services, under Contract No. 75N93021C00015. This work was supported in part by institutional funds from the University of Pennsylvania and NIH U19AI082630 (S.E.H. and E.J.W.). E.M.A. was supported by the NIH Training in Virology T32 Program (T32-AI-007324). We thank J. Lurie, J. Embiid, J. Harris, and D. Blitzer for philanthropic support. We thank all members of the Penn COVID-19 Sample Processing Unit. We would like to thank David Anderson for assistance with the graphical abstract. The contents of this publication are solely the responsibility of the authors and do not necessarily represent the official views of the NIH.

## AUTHOR CONTRIBUTIONS

Serological Assays, E.M.A., T.E., E.C.G, and M.J.B.,

Data Analyses, E.M.A.

Cohort Studies and Sample Processing, S.G., R.R.G, M.M.P., S.A.A., D.M., D.D., D.F., A.B., J.W., J.M., S.D., the UPenn COVID Processing Unit, and A.R.G.

Manuscript Writing, E.M.A., and S.E.H.;

Supervision, I.F., D.J.R, E.J.W., and S.E.H.;

Funding Acquisition, I.F., D.J.R, E.J.W., and S.E.H.

---

**Supplemental Table 1.**

|  | Infection | Vaccination | p-value |
| --- | --- | --- | --- |
| Total (N) | 10 | 23 |  |
| Median Age in years [min-max] | 35 [25-49] | 38 [23-67] | 0.8735 |
| Number of Female [%] | 8 [80%] | 15 [65%] | 0.6822 |

